# Antigen-specific T cell responses following single and co-administration of tick-borne encephalitis, Japanese encephalitis, and yellow fever virus vaccines: Results from an open-label, non-randomized clinical trial-cohort

**DOI:** 10.1101/2024.11.14.24317320

**Authors:** David Wullimann, John Tyler Sandberg, Mira Akber, Marie Löfling, Sara Gredmark-Russ, Jakob Michaëlsson, Marcus Buggert, Kim Blom, Hans-Gustaf Ljunggren

## Abstract

**Background:** Flavivirus infections pose a significant global health burden, highlighting the need for safe and effective vaccination strategies. Co-administration of different vaccines, including licensed flavivirus vaccines, is commonly practiced providing protection against multiple pathogens while also saving time and reducing visits to healthcare units. However, how co-administration of different flavivirus vaccines *de facto* affects immunogenicity, particularly with respect to T cell responses, is only partially understood.

**Methods and findings:** Antigen-specific T cell responses were assessed in study participants enrolled in a previously conducted open-label, non-randomized clinical trial. In the trial, vaccines against tick-borne encephalitis virus (TBEV), Japanese encephalitis virus (JEV), or yellow fever virus (YFV) were administered either individually or concomitantly in different combinations in healthy study participants. Peripheral blood samples were collected before vaccination and at multiple time points afterward. To analyze antigen-specific CD4+ and CD8+ T cell responses, PBMCs were stimulated with overlapping peptide pools from TBEV, JEV, YFV, and Zika virus (ZIKV) envelope (E), capsid (C), and non-structural protein 5 (NS5) viral antigens. A flow cytometry-based activation-induced marker (AIM) assay was used to quantify antigen-specific T cell responses. The results revealed remarkably similar frequencies of CD4+ and CD8+ T cell responses, regardless of whether vaccines were administered individually or concomitantly. In addition, administering the vaccines in the same or different upper arms did not markedly affect T cell responses. Finally, no significant cross-reactivity was observed between TBEV, JEV, and YFV vaccines, nor with related ZIKV-specific antigens.

**Conclusions:** TBEV or JEV vaccines can be co-administered with the live attenuated YFV vaccine without any markedly altered antigen-specific CD4+ and CD8+ T cell responses to the respective flaviviruses. Additionally, the vaccines can be delivered in the same or different upper arms without any significant influence on the T cell response. From a broader perspective, these results provide valuable insights into the outcome of immune responses following simultaneous administration of different vaccines for different but related pathogens.

**Author summary:** *Why was this study done?:*

- The World Health Organization recently declared a global initiative to control arboviral diseases. Many of these are caused by pathogenic flaviviruses, most transmitted by mosquitos and other arthropod vectors such as ticks.
- Vaccination is a key intervention for diseases caused by flaviviruses.
- Co-administration of different vaccines, including currently licensed flavivirus vaccines, is commonly practiced.
- Co-administration of vaccines saves time and reduces the number of visits to healthcare facilities and vaccine clinics.
- Cellular immune responses have not been thoroughly evaluated upon co-administration of currently licensed flavivirus vaccines, including yellow fever virus (YFV), tick-borne encephalitis virus (TBEV), and Japanese encephalitis virus (JEV) vaccines.

*What did the researchers find?:*

- The magnitude and specificity of CD4+ and CD8+ T cell responses to virus-specific antigens remained largely unchanged by the concomitant delivery of the studied flavivirus vaccines.
- Concomitant delivery of vaccines in the same or different upper arms of the study participants had minimal impact on CD4+ and CD8+ T cell responses.
- The studied vaccines maintained distinct CD4+ and CD8+ T cell reactivity across their respective viral antigens without generating any significantly detectable cross-reactivity to each other or ZIKV-antigens.

*What do these findings mean?:*

- Along with recently published data from the present study cohort, co- administration of three commonly used current licensed flavivirus vaccines is feasible without increasing the risk of adverse events or significantly affecting the development of either neutralizing antibodies (nAbs) or T cell responses against the respective viral antigens.

## Introduction

Yellow fever virus (YFV), tick-borne encephalitis virus (TBEV), and Japanese encephalitis virus (JEV) all belong to the *Flaviviridae* family of enveloped positive-sense RNA viruses [1]. These viruses pose major global health challenges, affecting both the inhabitants of endemic areas (around one-third of the world’s population) and travelers to these regions [2–4]. Infections with these viruses are associated with significant morbidity and numerous fatalities each year [5, 6]. No specific treatment for the diseases exists, though the development of antiviral drug candidates in the field is an active area of research [7].

A range of licensed flavivirus vaccines are currently available, including vaccines against YFV, TBEV, and JEV [8, 9]. These are frequently used to prevent disease among residents of endemic areas and travelers visiting these regions. In addition, several newer vaccines have been successful in late-phase clinical trials [10]. Safe and effective vaccine administration strategies are crucial [6]. While rigorous clinical trials have assessed the safety and immunogenicity of most currently approved vaccines, more limited data exist regarding administration strategies and the interactions between different flavivirus vaccines including the currently studied live attenuated YFV vaccine as well as inactivated TBEV and JEV vaccines [11].

To address this knowledge gap, we conducted an open-label, non-randomized clinical trial to assess the safety and serological response following concomitant delivery of commonly used licensed flavivirus vaccines [12]. The trial cohort included healthy adult volunteers who were vaccinated with either YFV and TBEV vaccines, YFV and JEV vaccines, or with the three respective vaccines alone. Half of the co-vaccinated participants received vaccines in the same upper arm, while the other half received the vaccines in different upper arms. Blood samples were collected before vaccination and at multiple time points afterward. Safety, as well as binding and nAb responses, were evaluated in detail [12]. However, the study did not assess antigen-specific T cell responses, an important part of the flavivirus immune response and induction of protective immunity [13].

In this study, we stimulated cryopreserved PBMCs from the above-mentioned clinical trial cohort with overlapping sets of peptides from TBEV, JEV, YFV, and ZIKV to identify antigen-specific CD4+ and CD8+ T cells. Stimulated PBMCs were assessed with an activation-induced marker (AIM) assay to determine the total T-cell response to major structural and non-structural flavivirus proteins, and to assess cross-reactivity.

Collectively, the present study provides insights into antigen-specific T cell responses following vaccination with different types of flavivirus vaccines, whether the vaccines were administered individually or concomitantly delivered to study subjects. From a broader perspective, the present results add further insight into the vaccine-induced immune responses that protect against diseases caused by flaviviruses.

## Methods

### Cohort and clinical samples

The PBMC samples used in this study are based on a completed clinical trial, referred to below. The clinical trial was designed as an open label, non-randomized academic (non-Pharma sponsored) trial [12]. It was conducted to assess safety and serological responses to three currently licensed flavivirus vaccines administered either as single agents or concomitantly delivered. Inclusion criteria allowed the participation of healthy volunteers,18 to 55 years of age, who sought protection from TBEV, JEV, or YFV or two of the three viruses (YFV and TBEV or YFV and TBEV). Participants were required to meet preset inclusion and exclusion criteria. All study volunteers had to sign informed consent documents in line with the ethical approval and clinical trial protocol. The trial was approved by the Stockholm Local Regional Ethical Committee (2017/1433-31/1) and the Swedish Medical Products Agency (5.1-2017-52376). It was registered and later on reported to the European database (EudraCT 2017-002137-32).

### Vaccines

Study subjects included in the clinical trial were vaccinated with the following flavivirus vaccines: Stamaril (Sanofi), live attenuated YFV 17D strain produced in pathogen-free chick embryo cells; IXIARO (Valneva), an inactivated, alum-adjuvanted, Vero cell-derived vaccine based on JEV strain SA14-14-2; FSME IMMUN (Pfizer), an inactivated, alum-adjuvanted, chick embryo cell-derived vaccine based on the TBEV Neudörfl strain [12]. All vaccines were obtained from Apoteket AB, Karolinska University Hospital, Solna, Sweden. Vaccinations were given in accordance with the clinical trial protocol and good clinical practice (GCP), with intervals for primary vaccinations as recommended in FASS (Pharmaceutical Specialties in Sweden): FSME IMMUN, three doses 0-, 1- and 5-month intervals; IXIARO, two doses, 0- and 1-month interval; Stamaril, one dose. For cohorts A1, A2, B1, B2, C, D, and E (for cohort description, see below), blood and serum were sampled before each vaccination and then on day 7 and 14 after each vaccination. Blood and serum were also sampled 30 days after the last vaccination (Fig 1A).

**Fig 1.**
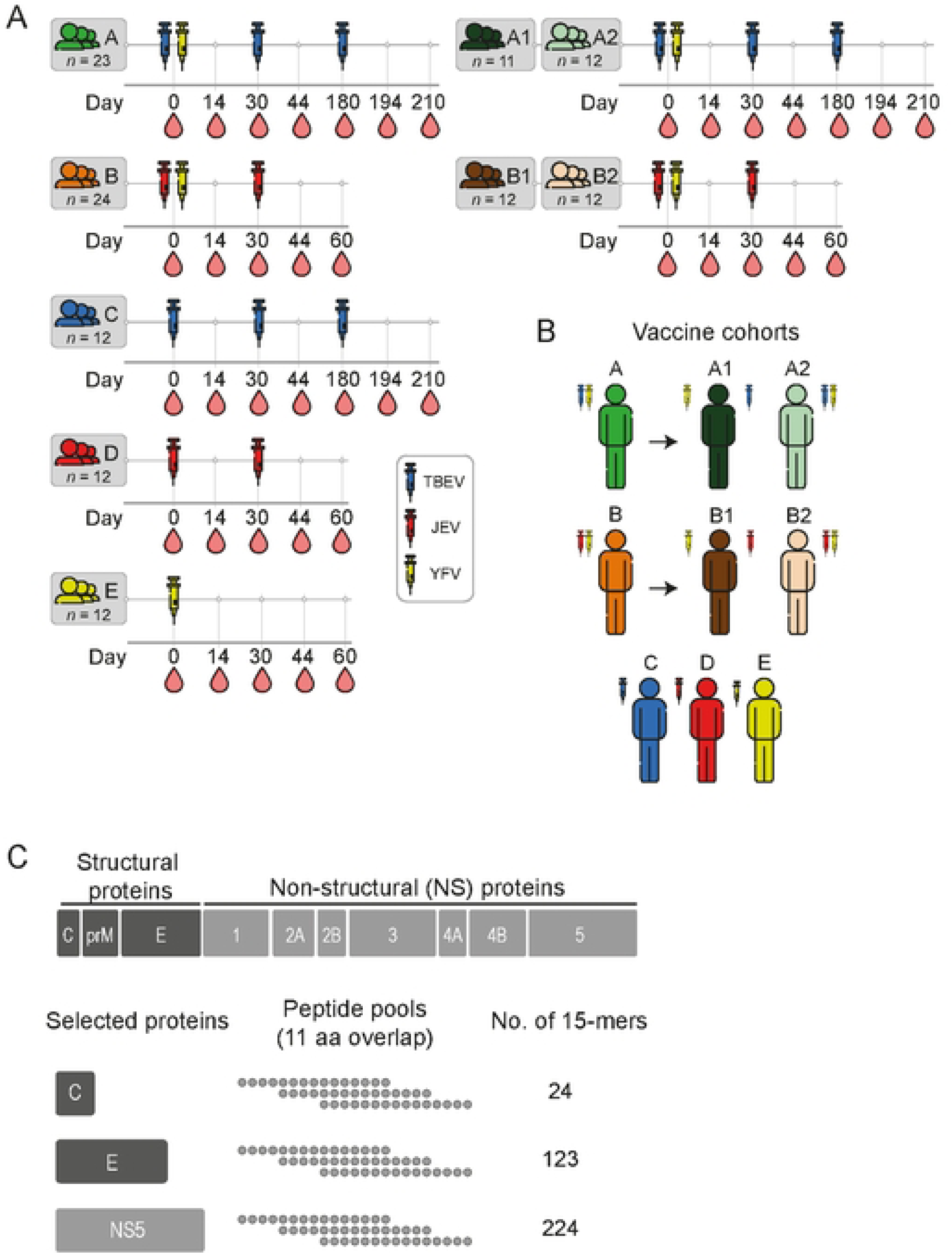
Overview of the vaccine cohorts **(A)** Blood sampling schedule and **(B)** administration sites for all vaccine cohorts included in the study. The number (*n)* of study participants in each vaccine cohort is written and applies to all subsequent Figs. **(C)** Out of the three structural and seven non-structural proteins encoded by the flavivirus genome, capsid (C), envelope (E), and non-structural protein-5 (NS5), were selected as targets for overlapping peptide pool- generation. Peptides, 15-mers with 11 amino acid (aa) overlap, were pooled for the respective protein and the number of peptides for each pool indicated.

### Study participants and cohorts

The clinical trial was initially designed to include 140 healthy volunteers [12]. Forty study participants were to receive both YFV and TBEV vaccines (cohort A). Twenty of these participants were to receive the vaccines in different upper arms (sub-cohort A1), and the other 20 in the same upper arm (sub-cohort A2). The next 40 study participants were to receive both YFV and JEV vaccines (cohort B). Similarly, 20 of these participants were to receive the vaccines in different upper arms (sub-cohort B1), and the other 20 in the same upper arm (sub-cohort B2). The remaining three cohorts consisted of 20 study participants per cohort and received either TBEV vaccine (cohort C), JEV vaccine (cohort D), or YFV vaccine (cohort E). Upon initiation of the clinical trial, a total of 161 healthy volunteers were screened for enrollment. 145 study participants were found eligible and enrolled in the trial. Among them, 43 participants were assigned to cohort A (A1, 23 participants; A2, 20 participants), 42 participants to cohort B (B1, 21 participants; B2, 21 participants), and 20 participants each to cohorts C, D, and E. Following enrollment three study participants dropped out from Cohort A1 and one study participant dropped out from each of cohorts B1, B2, and C. Hence, 139 study participants in total completed the trial. Throughout the trial, there were 13 missed visits out of a planned total of 1,150 visits [12].

### Viral strain sequences and alignments

The viral vaccine strains used for multiple sequence alignments and peptide pools were obtained from UniProt; TBEV (Neudörfl, P14336), JEV (SA14-14-2, P27395), YFV (YFV- 17D, P03314) and ZIKV (MR766, Q32ZE1). To determine percentage of homology between the different flaviviruses, multiple sequence alignment was performed with Clustal Omega [14].

### Peptides

Individual overlapping peptides consisting of 15-mers (11 amino acids overlap) were synthesized as crude material and delivered solubilized in 100% DMSO at 20 mg/ml (Merck). Each peptide for TBEV, JEV, and YFV as well as for ZIKV (sequences corresponding to viral strain mentioned above) E, C, and NS5 antigens was pooled and diluted in phosphate-buffered saline (PBS)to 20% DMSO. For a full list of peptides, see S1 Data set.

### Activation-induced marker (AIM) assay

To assess antigen-specific CD4+ and CD8+ T cell responses, a well-established AIM assay was used with slight modifications [15]. Briefly, PBMCs from study subjects (n=84) were thawed and resuspended in RPMI 1640 (Cytvia) supplemented with 10% Fetal bovine serum (Sigma-Aldrich), 1% Penicillin-streptomycin (Cytvia) and 1% L-glutamine (Sigma-Aldrich). One million cells per well were cultured in U-bottom 96-well plates (Corning) for 3 h at 37°C to rest. After resting, cells were stimulated with a mix containing the appropriate peptide pool (0.5 µg/ml), unconjugated anti-CD40, and anti-CXCR5 Abs. After a 14 h incubation at 37°C and 5% CO2, the plate was centrifuged at 1,800 rpm for 3 min, and the supernatant was removed. The cells were transferred to V-bottom 96-well plates (ThermoFischer Scientific) and washed with PBS. Cells were stained for viability for 10 min, incubated with anti-CCR7 and anti-CX3CR1 at 37°C for 10 min, followed by the remaining fluorophore-conjugated Abs which were mixed with Brilliant Violet Buffer Plus (BD Biosciences) and FACS buffer (PBS with 2% FBS and 2 mM ethylenediaminetetraacetic acid [EDTA]) for 30 min at room temperature. All Abs used are listed in S1 Table. Stained cells were washed, fixed with FoxP3/Transcription Factor Buffer Set (ThermoFischer Scientific), and acquired on a BD FACS Symphony A3 (BD Biosciences). Samples were analyzed with FlowJo v.10 (TreeStar, Inc.) and quality control was performed with the flowAI algorithm [16]. Frequencies of CD69+CD40L+ of CD4+ T cells and CD69+41BB+ of CD8+ T cells were subtracted with each respective DMSO control. Negative values after subtraction were set to 0.001 as the limit of detection (LOD) to allow for logarithmic scale display in graphs. For each PBMC sample, DMSO at the same concentration as in the respective peptide pools was used as a negative control and 1μg/ml SEB was used as a positive control.

### Statistics

Statistical analysis was performed in Python (version 3.12.4) using SciPy (version 1.11.4) and scikit-posthocs (version 0.9.0). All plots and statistical annotations were generated using pandas (version 1.5.3), numpy (version 1.26.3), seaborn (version 0.13.2) and matplotlib (version 3.7.1) through custom scripts. Data distributions from flow cytometry experiments were determined to be non-parametric using diagnostic plots and the Shapiro-Wilk test for normality. For unpaired data, comparisons between three or more groups were analysed using the Kruskal-Wallis’s test followed by the Dunn’s post hoc test. For unpaired data between two groups, the Mann-Whitney rank test was used. Paired data were analysed using the Friedman test followed by Siegel and Castellan’s All-Pairs Comparisons post hoc test. Post hoc test results were corrected using the Bonferroni method (adjusted alpha = 0.002 for comparisons between cohorts A and C, and adjusted alpha = 0.005 cohorts B, D and E). All tests were two-sided, with statistically significance set at p <0.05. The adjusted p-value is reported in figures when correction for multiple comparisons was applied.

## Results

### Overview of vaccine cohorts

Out of the total 139 study subjects, clinical samples from 84 study subjects (12 randomly selected from each of the respective seven study cohorts A1, A2, B1, B2, C, D, and E) were selected for inclusion in the present study (Fig 1A and B). One study participant from cohort A1 was later excluded from all data analyses due to the detection of nAbs against TBEV at day 0, leaving 83 study subjects remaining in the present study. PBMCs were stimulated with overlapping peptide pools consisting of 15- mers spanning the entire length of the two structural proteins envelope (E) and capsid (C), as well as the large and conserved non-structural protein-5 (NS5) (Fig 1C).

### Kinetics of the antigen-specific CD4+ and CD8+ T cell vaccine response

First, we assessed antigen-specific T cell responses in the three cohorts that received TBEV (cohort C), JEV (cohort D), and YFV (cohort E) vaccines, respectively (Fig 2). CD4+ and CD8+ T cell responses were evaluated using an AIM assay (Fig 2A and S1 Fig).

**Fig 2.**
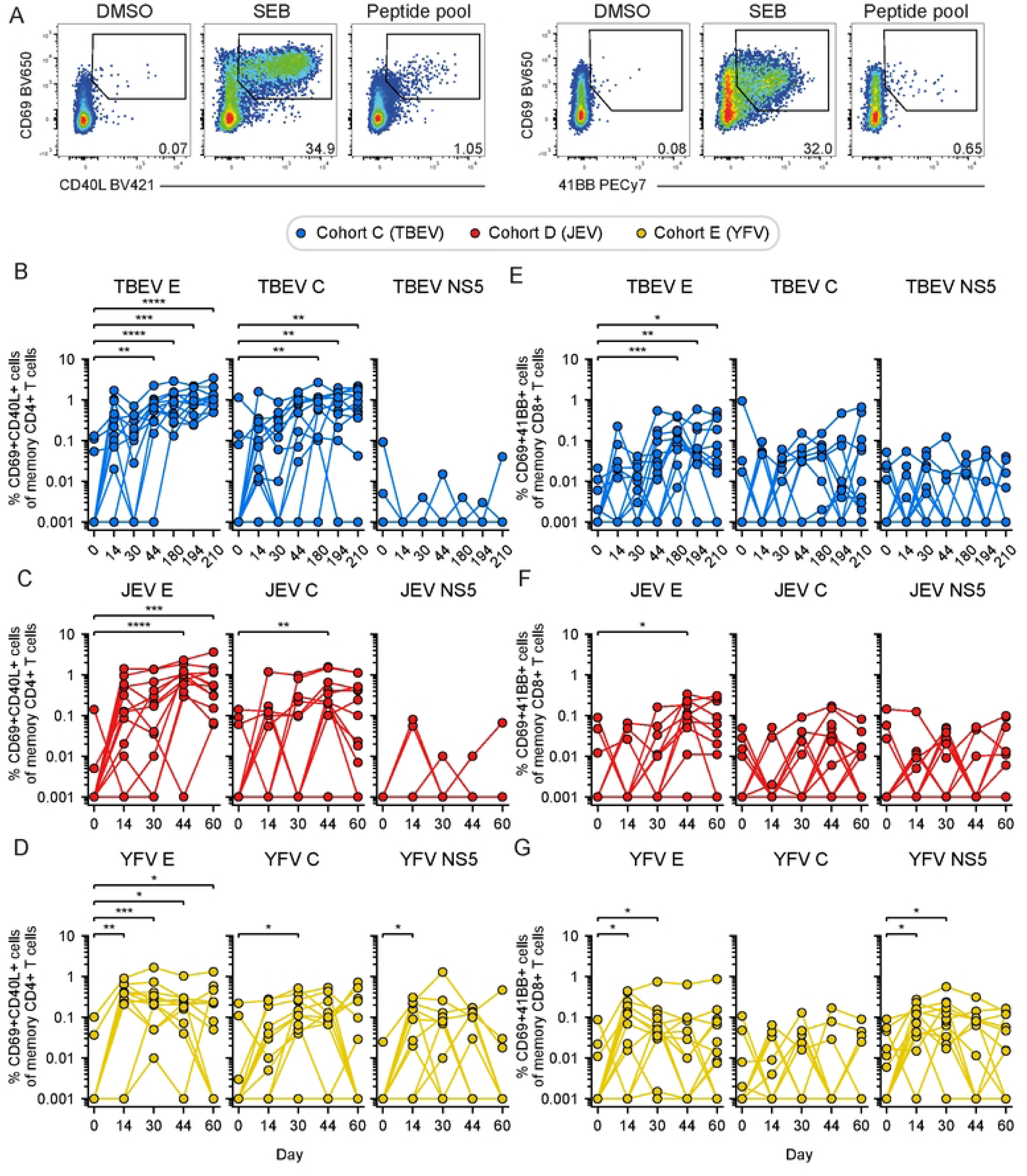
Kinetics of the antigen-specific CD4+ and CD8+ T cell response in study participants vaccinated against TBEV, JEV, and YFV. **(A)** Representative flow cytometry gating for the AIM assay used to identify antigen-specific CD4+ T cells (CD69+CD40L+) and antigen-specific CD8+ T cells (CD69+41BB+) after PBMC stimulation with DMSO (negative control), SEB (positive control), and peptide pool (E, C or NS5). **(B-D)** Frequency of TBEV-, JEV- and YFV-specific E, C, and NS5 CD4+ T cells in the **(B)** TBEV (cohort C), **(C)** JEV (cohort D) and **(D)** YFV vaccinated (cohort E), respectively. **(E-G)** Frequency of TBEV-, JEV- and YFV-specific E, C, and NS5 CD8+ T cells in the **(E)** TBEV (cohort C), **(F)** JEV (cohort D) and **(G)** YFV vaccinated (cohort E), respectively. **(D-G)** Statistical analysis assessed by Friedman test, with Siegel and Castellan’s All-Pairs Comparisons post hoc tests with Bonferroni correction. Significant results are shown, where: *p ≤ 0.05, **p ≤ 0.01, ***p ≤ 0.001, ****p ≤ 0.0001.

Significantly higher TBEV E- and C-specific CD4+ T cell responses were observed in the TBEV vaccinated cohort over time compared to baseline (Fig 2B). Similarly, significantly higher JEV E- and C-specific CD4+ T cell responses were observed over time compared to baseline (Fig 2C). In contrast, no detectable NS5-specific CD4+ T cell response was observed after vaccination with either TBEV or JEV vaccines (Fig 2B and C). Following YFV vaccination, YFV-specific CD4+ T cell responses were observed against both E, C, and NS5 (Fig 2D). The overall activation of antigen-specific CD8+ T cells was lower in participants who received the inactivated vaccines against TBEV (Fig 2E) and JEV (Fig 2F), while significant E- and NS5-specific CD8+ T cell responses were detected in participants who received the live attenuated YFV vaccine (Fig 2G). In summary, the AIM assay effectively captured the overall kinetics of the antigen-specific T cell response following vaccination with inactivated TBEV and JEV vaccines as well as with the live attenuated YFV flavivirus vaccine.

### Co-vaccination retains the antigen-specific T cell response

Overall, the CD4+ and CD8+ T cell response against E epitopes was the most dominant antigen-specific T cell response across all vaccine cohorts (Fig 2). To evaluate the influence of concomitant delivery of vaccines on the antigen-specific T cell response, we compared the results from the single vaccinated TBEV, JEV, and YFV cohorts (cohorts C, D, and E) with those from the two co-vaccinated cohorts (cohorts A and B); the latter co-vaccinated with TBEV and YFV (TBEV+JEV) as well as JEV and YFV (JEV+YFV) vaccines, respectively (Fig 1). Initial analyses revealed similar kinetics in terms of CD4+ and CD8+ T cell responses in cohort A and cohort B, respectively (S2 Fig). Importantly, overall, no major differences in the magnitude of TBEV or JEV E-specific CD4+ T cell responses were observed when comparing the respective single and co-vaccinated cohorts (Fig 3A and B). Similar response patterns, though generally weaker than the CD4+ T cell responses, were observed among TBEV or JEV E-specific CD8+ T cells (Fig 3C and D). Additionally, we investigated whether the YFV response was influenced by co-vaccination with TBEV or JEV vaccines. Overall, the observed YFV-specific T cell responses were similar (S3A Fig) or lower (S3B-D Fig) in the single vaccinated compared to the co-vaccinated cohorts, though the significance of this finding is not fully clear (see Discussion).

**Fig 3.**
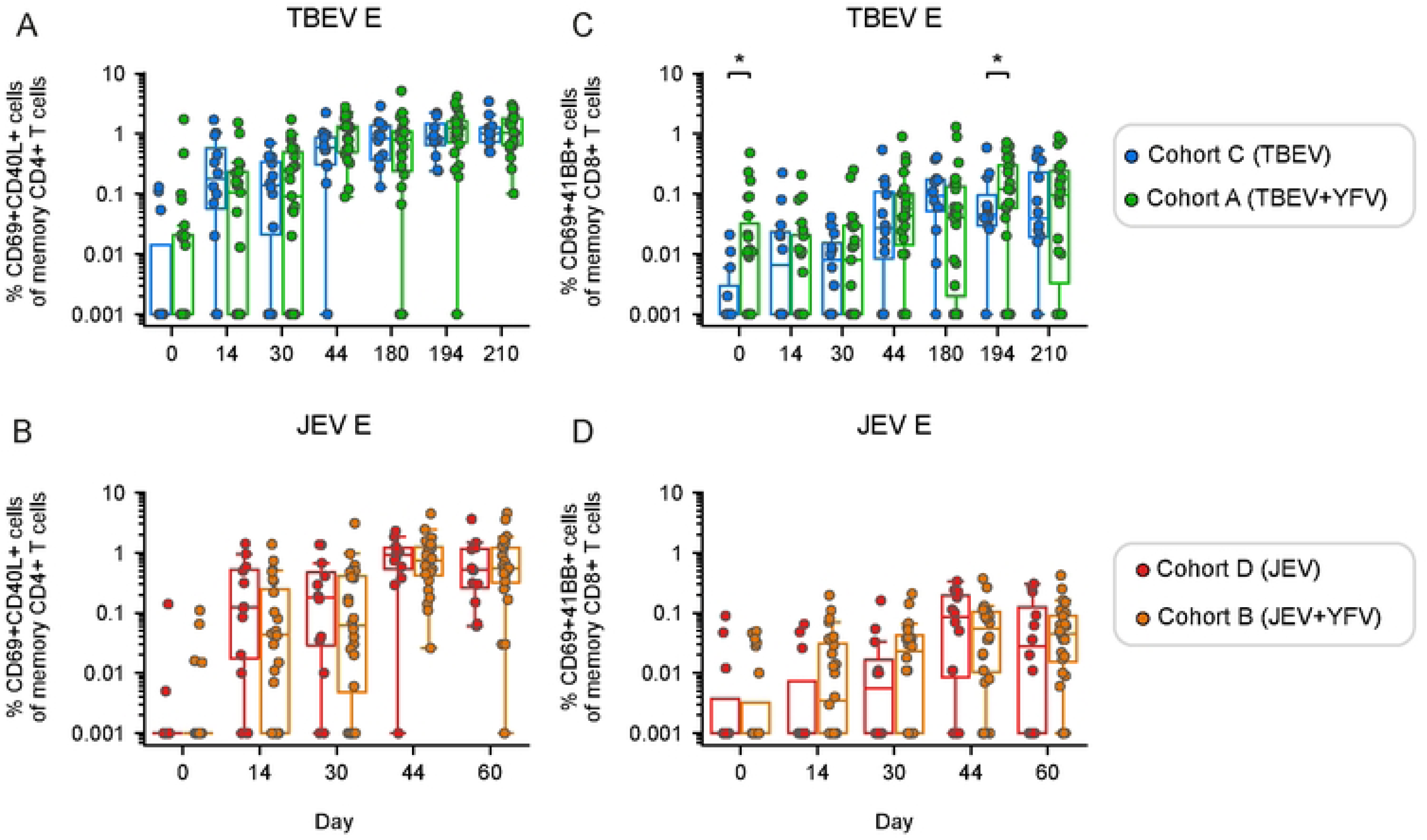
Influence of co-vaccination on antigen-specific CD4+ and CD8+ T cell responses. **(A)** Frequency of TBEV E-specific CD4+ T cells between study participants from cohort C (TBEV) and cohort A (TBEV+YFV) and **(B)** frequency of JEV E-specific CD4+ T cells between cohort D (JEV) and cohort B (JEV+YFV). **(C)** Frequency of TBEV E-specific CD8+ T cells between cohort C and cohort A and **(D)** frequency of JEV E-specific CD8+ T cells between cohort D and cohort B. **(A-D)** Statistical analysis assessed by Mann-Whitney rank test at each time point. Distribution of data points is determined by boxplot with median and 25^th^-75^th^ percentiles; whiskers are drawn from 1.5 times the IQR. Significant results are shown, where: *p ≤ 0.05, **p ≤ 0.01, ***p ≤ 0.001, ****p ≤ 0.0001.

### Co-vaccination in the same or different upper arm does not impact the antigen-specific T cell response

To determine whether concomitant delivery of the TBEV or JEV vaccines with the YFV vaccine affects immunogenicity depending on whether the vaccines were administered in the same or different upper arms, we analyzed specific subgroups of the co-vaccinated TBEV+YFV (cohort A1 and A2) and JEV+YFV (cohort B1 and B2) cohorts (Fig 1). The frequency of TBEV E-specific CD4+ T cells was overall similar when the two TBEV+YFV subgroups were compared most apparent when evaluating results at the peak of responses and/or later timepoints (Fig 4A). Likewise, the frequency of JEV E-specific CD4+ T cells was overall similar when the two JEV+YFV subgroups were compared (Fig 4B). Overall, analysis of the CD8+ T cell responses yielded similar results in terms of comparisons between groups vaccinated in the same or different upper arms (Fig 4C and Fig 4D), Additionally, the CD8+ T cell responses were overall weaker than CD4+ T cell responses, in line with our previous results (Figs 2 and 3). Similar patterns were observed for the YFV E-specific CD4+ and CD8+ T cell response, comparing corresponding groups (S4 Fig). Taken together, these data indicate that multiple immunizations in the same or different upper arms do not markedly influence the outcome the generation of CD4+ and CD8+ T cell responses.

**Fig 4.**
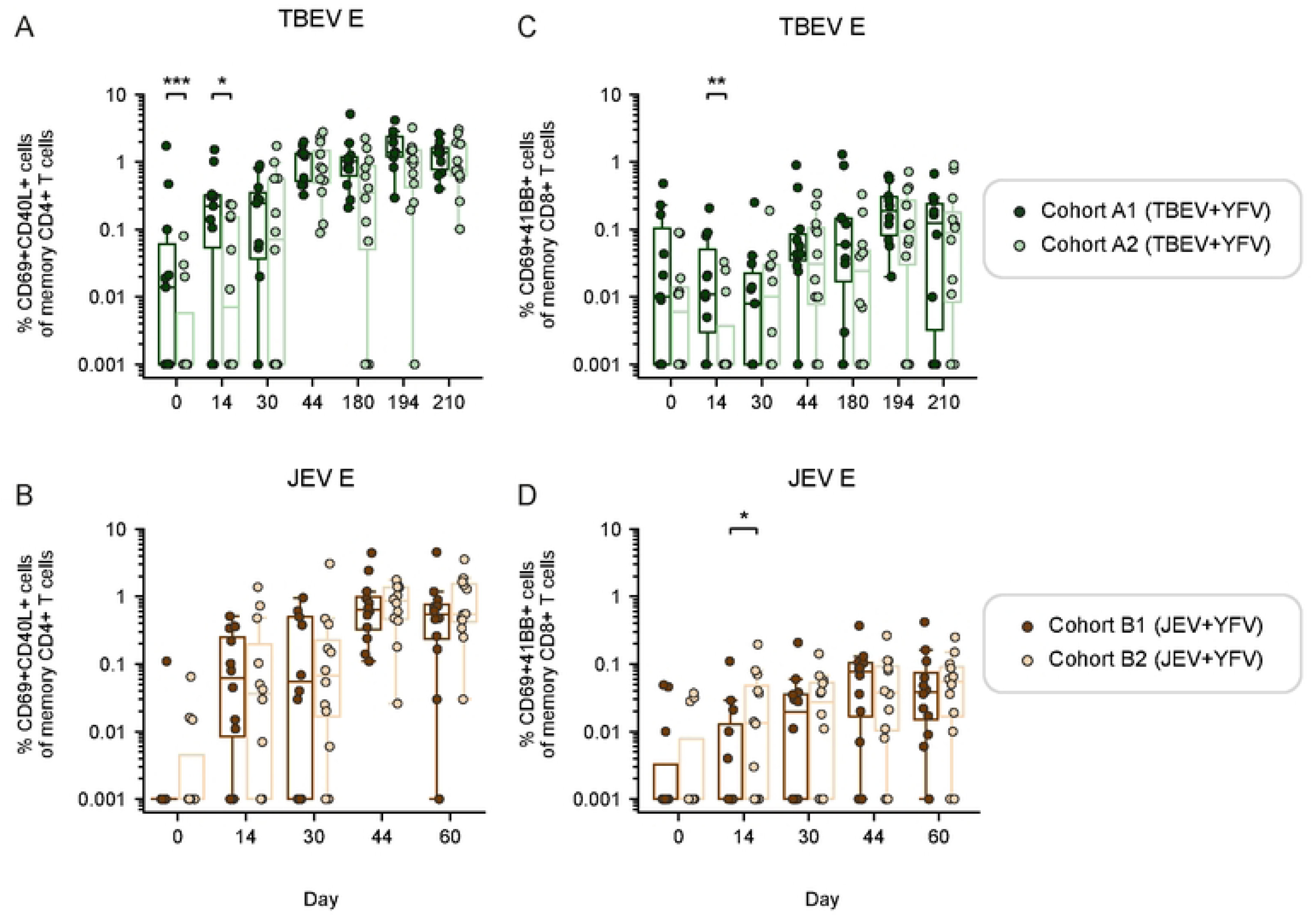
Influence of co-vaccination on antigen-specific T cell responses upon vaccination in the same or different upper arms. **(A)** Frequency of TBEV E-specific CD4+ T cells compared between cohort A1 and A2. **(B)** Frequency of JEV E-specific CD4+ T cells compared between cohort B1 and B2. **(C)** Frequency of TBEV E-specific CD8+ T cells compared between cohort A1 and A2. **(D)** Frequency of JEV E-specific CD8+ T cells compared between cohort B1 and B2. **(A-D)** Statistical analysis assessed by Mann-Whitney rank test at each time point. Distribution of data points is determined by boxplot with median and 25^th^-75^th^ percentiles; whiskers are drawn from 1.5 times the IQR. Significant results are shown, where: *p ≤ 0.05, **p ≤ 0.01, ***p ≤ 0.001, ****p ≤ 0.0001.

### Effector memory T cells (TEM) constitute the main memory phenotype of antigen-specific T cells

We further investigated the phenotypical characteristics of the activated CD4+ and CD8+ T cells in the vaccine cohorts. The memory phenotypes studied for CD4+ and CD8+ T cells included central memory (TCM, CCR7+CD45RA-), effector memory (TEM, CCR7-CD45RA-), effector memory T cells re-expresses CD45RA (TEMRA, CCR7- CD45RA+), as well as CD4+ T follicular helper cells (Tfh, CXCR5+). Across the studied cell populations, we observed an increase in the frequency of CD4+ TEM cells following vaccination in the study cohorts (Fig 5 and S5 Fig). As a result, remaining populations, including Tfh cells, decreased in frequencies over time following vaccination (S5 Fig). The expansion of CD4+ TEM cells follow as expected the initial expansion of AIM+ populations towards specific viral proteins as shown in Figs 2-4. These observations are in line with the notion that many of the above-mentioned CD4+ T cell populations, including TCM and Tfh, may migrate to lymphoid tissues upon antigen activation [17].

**Fig 5.**
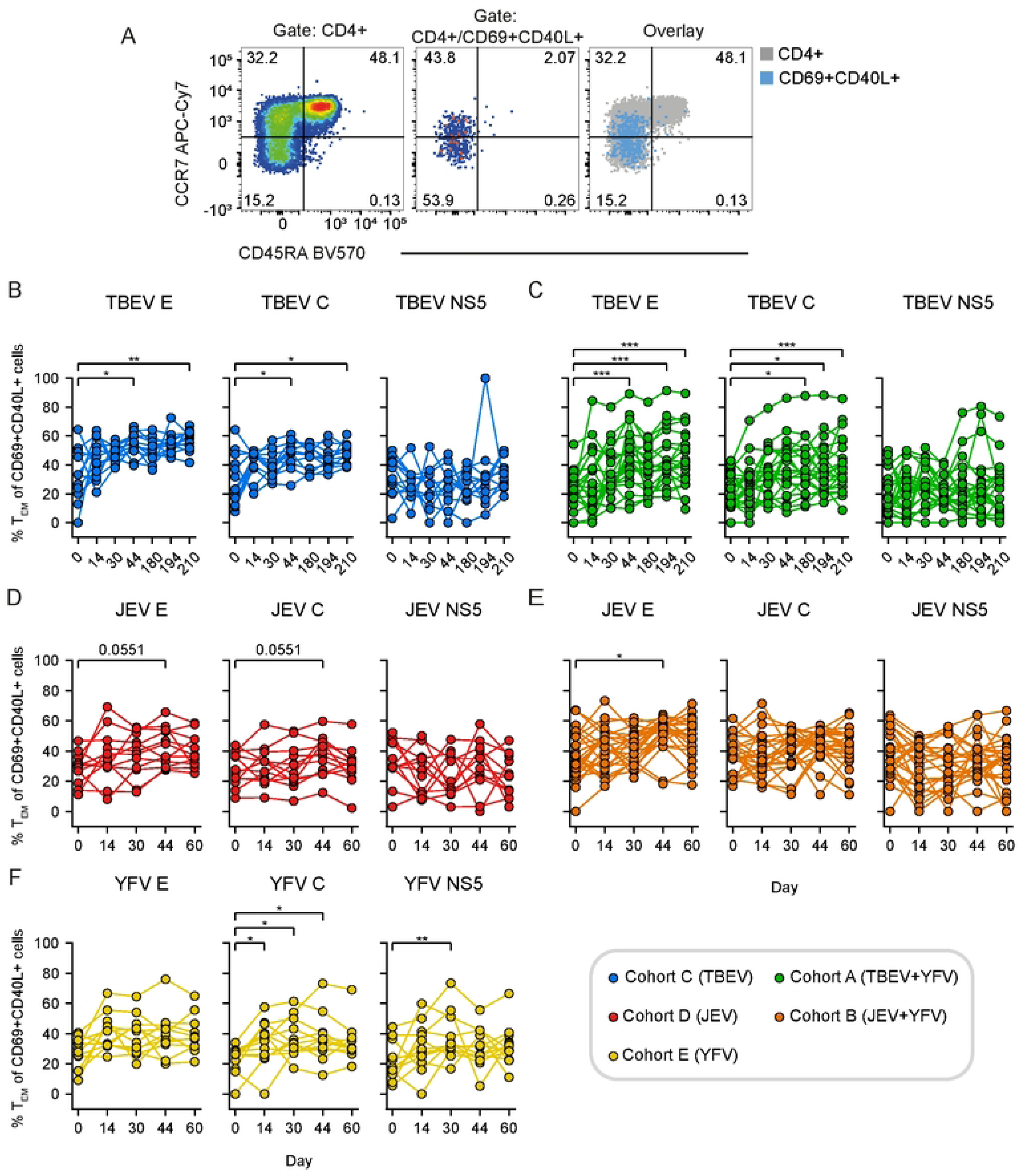
Kinetics of the antigen-specific CD4+ memory effector T cell (TEM) responses. **(A)** Representative flow cytometry gating of memory CD4+ T cells based on CD45RA and CCR7 expression within bulk CD4+ T cells (left panel) and within CD69+CD40L+ of CD4+ T cells after peptide stimulation (right panel) . **(B)** Frequency of TBEV E-, C- and NS5- specific CD4+ TEM cells in TBEV vaccinated (cohort C). **(C)** Frequency of TBEV E-, C- and NS5-specific CD4+ TEM cells in YFV+TBEV vaccinated (cohort A). **(D)** Frequency of JEV E-, C- and NS5-specific CD4+ TEM cells in JEV vaccinated (cohort D). **(E)** Frequency of JEV E-, C- and NS5-specific CD4+ TEM cells in YFV+JEV vaccinated (cohort B). **(F)** Frequency of YFV E-, C- and NS5-specific CD4+ TEM cells in YFV vaccinated (cohort E). **(B-F)** Statistical analysis assessed by Friedman test, with Siegel and Castellan’s All-Pairs Comparisons post hoc tests with Bonferroni correction. Significant results are shown, where: *p ≤ 0.05, **p ≤ 0.01, ***p ≤ 0.001, ****p ≤ 0.0001.

### The vaccine-induced T cell response showed limited cross-reactivity towards other flavivirus antigens

Because similarities in amino acid sequences in viral proteins can drive cross-reactive immune responses between different viruses, we conducted multiple amino acid sequence alignments of the flaviviruses included in this study to characterize their overall sequence similarity. The E protein has 41-44% similarity when compared between TBEV, JEV, and YFV. The C protein has the lowest similarity, ranging from 24-31% similarity, while the highest sequence similarity was seen within the NS5 protein with 61-63% similarity (Fig 6A). To address the extent to which T cell cross-reactivity among the flaviviruses studied exists, we compared TBEV, JEV, and YFV E-, C-, and NS5- specific CD4+ and CD8+ T cell responses in all vaccine cohorts. With respect to E responses, we observed very limited cross-reactive T cell responses across all vaccine cohorts, including CD4+ (Fig 6B and C) and CD8+ T cell responses (Fig 6D and E). Similar results were also observed for CD4+ and CD8+ T cell responses against C (S6 Fig) and NS5 (S7 Fig). The results were further corroborated by analyses of increases in the median response on peak response days against the respective E, C, and NS5 antigens (S8-13 Figs). Taken together, these data demonstrate that the present flavivirus vaccines induce, at most, very limited cross-reactive T cell responses against each other.

**Fig 6.**
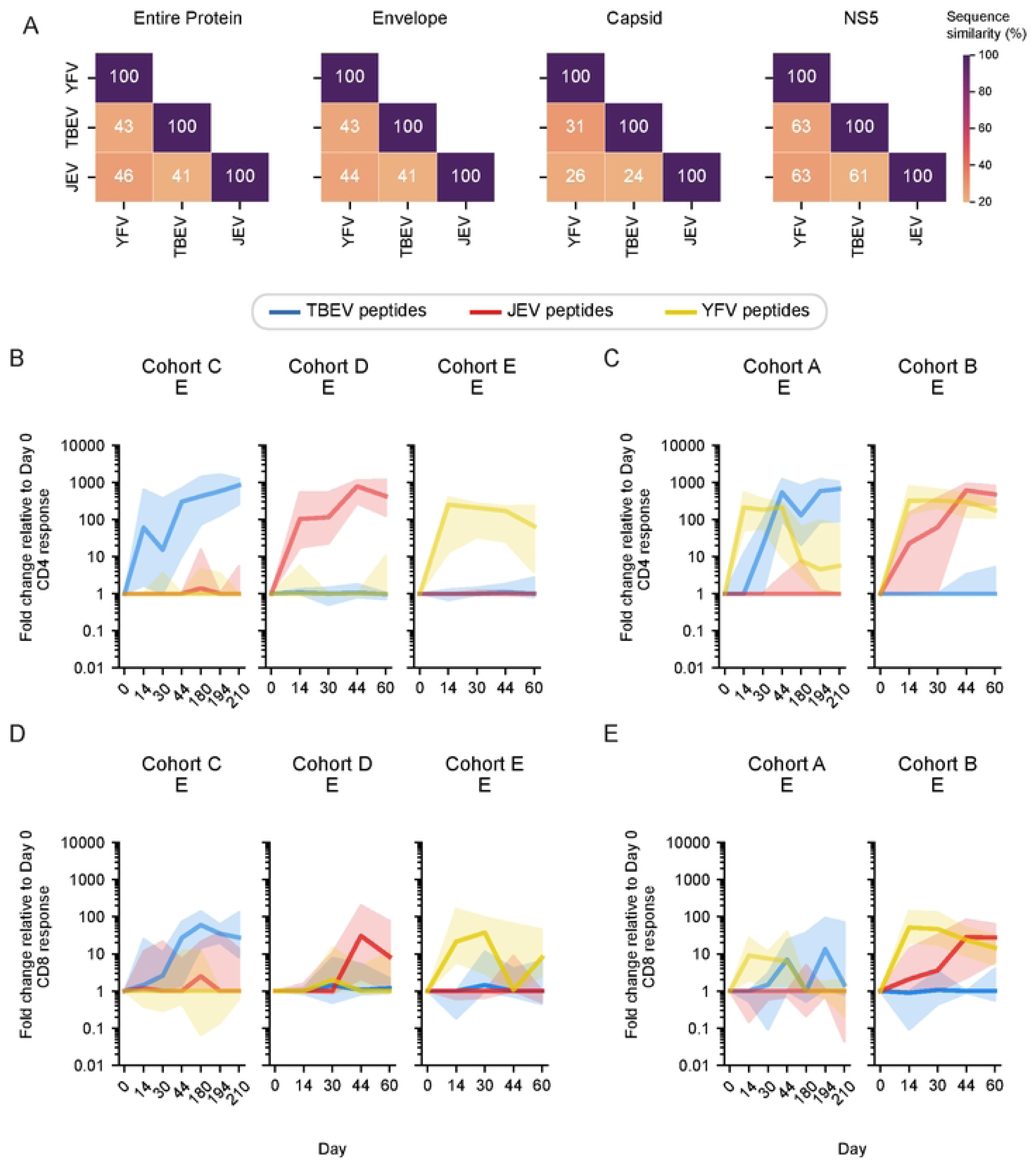
Cross-reactive E-specific T cell responses in the vaccine cohorts. **(A)** Sequence alignments between TBEV, JEV, and YFV. Heatmaps are annotated according to percent of amino acid sequence similarity. **(B-C)** Frequency of TBEV, JEV, and YFV E-specific CD4+ T cells, and **(D-E)** TBEV, JEV, and YFV E-specific CD8+ T cells, in study participants from all vaccine cohorts expressed as fold change relative to day 0. Line plots show median and 95% confidence interval (shaded area).

### Limited cross-reactivity against Zika virus antigens

Zika virus (ZIKV) also possesses amino acid sequence similarities with the other flaviviruses studied here (Fig S14A). This led to us to assess potential cross-reactivity to ZIKV antigens following vaccination with the present studied vaccines, with a particular focus on studies of cross-reactive responses elicited by the YFV vaccine. To this end, we did not detect any marked vaccine-induced CD4+ or CD8+ T cell reactivity against the ZIKV antigens (E, C, and NS5) in any of the three YFV, YFV+TBEV, or YFV+JEV vaccinated cohorts (Fig S14B-G).

## Discussion

The present study presents findings from a non-randomized clinical trial cohort in which healthy study subjects received single or concomitant administrations of different flavivirus vaccines directed against TBEV, JEV, and YFV. Flow cytometry-based analysis revealed robust antigen-specific CD4+ and CD8+ T cell responses across all studied cohorts, with similar levels as single-vaccinated regardless of the concomitant vaccine-delivery or the site of administration. Additionally, the vaccines did not induce significant T cell cross-reactivity among each other or to ZIKV specific antigens.

Vaccines remain the most effective interventions for many flavivirus-induced diseases [8]. Because of the general knowledge gap in relation to safety and immunogenicity-related responses on concomitantly delivered vaccines [18], the recently conducted open label, non-randomized clinical trial was designed to provide safety and Ab immunogenicity-related data upon concomitant delivery of TBEV, JEV, and YFV vaccines [12]. However, the pivotal study did not address antigen-specific cellular immune responses. The present results complement the latter by showing that concomitant vaccine delivery does not markedly affect the generation of antigen-specific T cell responses, neither enhancing nor suppressing them. Additionally, immunization in the same or different upper arms did not largely impact the generation of antigen-specific T cell responses. The rationale for addressing the latter question was that local innate adjuvant-related effects from one vaccine might affect responses to the other vaccine, or that induction of specific immunity in the same local draining lymph nodes might skew the response towards one type of antigen over another. The recently reported binding and nAb-responses from the same study cohort [12] are in line with the above-mentioned conclusions. However, notably, we did observe an indication of an increased YFV-specific T cell response in the co-vaccinated study participants compared to the single YFV-vaccinated study participants (S3 Fig). Further research is needed to assess the validity of this finding in a larger study cohort.

The present study demonstrated marked CD4+ T cell responses in all vaccine cohorts, while CD8+ T cell responses were predominantly detected in the YFV vaccine recipients. It should be noted that the lower and more heterogenous detection of CD8+ T cells is not unexpected with stimulation using 15-mer peptides. These observations are in line with earlier studies on T cell reactivity to YFV vaccination [19–21] and to more limited studies published with respect to T cell responses to TBEV [22–24] and JEV [25] vaccination. In this context, it has been described that alum-adjuvanted flavivirus vaccines induce more of a Th2-biased response and, in this, may avoid a more, sometimes immunopathological, Th1 and/or CD8+ T cell response [8]. The present study also mapped CD4+ and CD8+ T cell responses specifically against structural and non-structural proteins from YFV, TBEV, JEV, and additionally ZIKV using overlapping peptide pools targeting the entire E, C, and NS5 proteins. While nAbs predominantly target the E protein [11, 26, 27], T cell responses can be directed against both structural and non-structural flavivirus proteins [13, 28]. TBEV and JEV inactivated vaccine recipients predominantly showed responses to E and C, while live attenuated YFV vaccine recipients developed responses against all antigens, where the highest reactivities detected were against the E and NS5 antigens. Consistent with these observations, previous studies have shown that inactivated vaccines, such as those against TBEV and JEV, do not generate efficient T cell responses specific to NS proteins, resulting in lower CD8+ T cell activation and reduced immunogenicity compared to the live attenuated YFV vaccine [22, 24].

Several studies have addressed the role of pre-existing immunity upon flavivirus vaccination [29–38]. For example, in a study by Lima and collaborators, pre-existing immunity from prior JEV vaccination, but not YFV vaccination, was found to influence the duration of the CD4+ T cell response following vaccination with an inactivated ZIKV vaccine [39]. In relation to this, our longitudinal TBEV- and JEV-vaccination and consecutive sampling strategy also allowed for investigations of the influence of pre-existing immunity from YFV vaccination on the T cell response to TBEV- and JEV-booster doses up to 210 and 60 days, respectively. Assessing the present data, we did not detect a significant enhancement or reduction in T cell responses towards TBEV or JEV epitopes in the YFV co-vaccinated groups, indicating that pre-existing immunity from YFV vaccination does not markedly influence the CD4+ and CD8+ T cell response to TBEV or JEV booster doses, at least not within the present study (rather short) period.

In relation to the discussion above, we also more specifically addressed potential T cell cross-reactivities between the vaccine-induced immune responses. In short, we found no evidence that vaccination with YFV, TBEV, or JEV alone or in combination resulted in any significant cross-reactive T cell response to viral antigens other than those specific to the vaccine used for vaccination. This included no detectable T cell responses against ZIKV virus epitopes. The present results align with those from a comprehensive study by Grifoni and colleagues [40]. In that study, vaccination with YFV-17D resulted in limited cross-reactive T cells against DENV, ZIKV, West Nile virus (WNV), and JEV antigens.

Additionally, cross-reactive T cells were predominately seen against 9 or 10-mers with a sequence similarity above 67%. In line with the findings above, the lack of cross-reactive T cell responses in the present study can be explained by the relatively long antigenic distance between YFV, JEV, and TBEV, which correspond to three different serocomplexes. Furthermore, the YFV serocomplex has the highest genetic distance from other flavivirus serocomplexes, where most of the conserved T cell epitopes are lost [41]. Epidemiological studies further support the present conclusions. For example, no protective association was found between YFV antibody titers and protection against serious adverse outcomes from ZIKV exposure in utero [42]. In this context, it is noted that vaccination against YFV early on following the ZIKV outbreak in 2015 was suggested as a possible means to limit the severe consequences of ZIKV infection in the absence of a ZIKV-specific vaccine [43]. However, it cannot be excluded that responses to other combinations of flavivirus vaccines or combinations of vaccines from other vaccine platforms than the present ones studied could result in cross-reactive T cell responses [44].

The relative strength of the present study is that data were collected from a *bona fide* prospective clinical trial cohort with the scientific and regulatory rigor inherent to this design, including independent study monitoring of original clinical trial data. It complements recently published clinical safety and serological data from the same study cohort addressing the effects of concomitant delivery of different flavivirus vaccines [12]. With respect to the limitations of this study, larger study groups could have given even more robust data. Deeper analyses on responding T cell populations could have given more detailed insights into other characteristics of responding T cell populations. Studies could have been performed also against other flavivirus antigens than E, C and NS5. The absence of screening for seropositivity to flavivirus exposure other than those the study subjects were vaccinated with prior to enrollment into the study is a relative limit. However, with respect to this, only two study participants with positive nAbs against TBEV and no one to JEV or YFV were identified among the entire cohort (n=139). One of these study subjects randomly picked for the present study had to be omitted from further analysis. Hence, the present study came to include 83 of the intended 84 study subjects.

In conclusion, the present study, together with data from the previous clinical trial [12], underscores the development of robust Ab and T cell responses elicited by flavivirus vaccinations. The two studies support the safety and efficacy of simultaneous vaccine administration strategies with the herein studied flavivirus vaccines. These findings provide valuable insights into the optimization of vaccination protocols, highlighting the compatibility of different vaccination regimens with minimal impact on immune response quality and safety profiles. This is particularly important as the world becomes increasingly susceptible to flavivirus-caused diseases as a consequence of climate change facilitating the geographical expansion of insect vectors, habitat, and urban environmental changes, as well as extensive global traveling [6, 8].

## Data Availability

All relevant data are within the manuscript and its Supporting Information files.

## Acknowledgments

We gratefully acknowledge the contributions of all study participants in the original clinical trial. We also extend our appreciation to the Karolinska Trial Alliance staff and research nurses for their exceptional organization of participant recruitment, scheduling, vaccination, and clinical sampling. Special thanks go to R. Varnaité, J. Emgård, N. Al-Tawil, L. Lindquist, J. Klingström and K. Loré for their dedicated efforts and contributions to the original clinical trial. We are grateful to Y. Gao for providing methodological advice. Additionally, we thank E. Alici and D. Calder for their support during the initial phase of this project.

## URL to website

https://ki.se/en/medh/hans-gustaf-ljunggren-group-immune-responses-to-human-virus-infections-and-cancer

## Abbreviations

Ab: antibody
AE: adverse events
AIM: activation-induced marker
C: capsid
DENV: dengue virus
DMSO: Dimethyl sulfoxide
E: envelope
JEV: Japanese encephalitis virus
nAb: neutralizing antibody
NS5: non-structural protein 5
PBMC: peripheral blood mononuclear cells
SEB: Staphylococcal enterotoxin B
TBEV: tick-borne encephalitis virus
YFV: yellow fever virus
WNV: West Nile virus
ZIKV: Zika virus

## Supporting information

S1 Fig. Flow cytometry gating strategy for identification of AIM markers and memory phenotypes.

Lymphocytes were gated followed by subsequent singlet gating. Cells were then gated as CD3^+^ and Dump^-^ (dead cells, CD19+ and CD14+). T cells were subdivided as either CD4^+^ or CD8^+^ cells. CD4+ T follicular helper cells were identified as CXCR5+. CD4+ and CD8+ (bulk) memory cell phenotypes were defined by CD45RA and CCR7 expression. CD4+ and CD8+ AIM+ cells were gated on bulk and non-naive cells (CCR7+CD45RA+). Antigen-specific T cells were identified with AIM markers, where antigen-specific CD4+ T cells were defined by CD69^+^CD40L^+^ and antigen-specific CD8+ T cells as CD69^+^41BB^+^.

S2 Fig. Kinetics of the antigen-specific T cell response in cohort A and B.

**(A-B)** Frequency of TBEV- and JEV-specific E, C, and NS5 CD4+ T cells in the **(A)** TBEV+YFV (cohort A) and **(B)** JEV+YFV (cohort B), respectively. **(C-D)** Frequency of TBEV- and JEV-specific E, C, and NS5 CD8+ T cells in the **(C)** TBEV+YFV (cohort A) and **(D)** JEV+YFV (cohort B), respectively. **(A-D)** Statistical analysis assessed by Friedman test, with Siegel and Castellan’s All-Pairs Comparisons post hoc tests with Bonferroni correction. Significant results are shown, where: *p ≤ 0.05, **p ≤ 0.01, ***p ≤ 0.001, ****p ≤ 0.0001.

S3 Fig. Influence of co-vaccination on the YFV-specific T cell response.

**(A)** Frequency of YFV E-specific CD4+ T cells between study participants from cohort E (YFV) and cohort A (TBEV+YFV) and **(B)** frequency of YFV E-specific CD4+ T cells between study participants from cohort E and cohort B (JEV+YFV). **(C)** Frequency of YFV E-specific CD8+ T cells between study participants from cohort E and cohort A and **(D)** frequency of YFV E-specific CD8+ T cells between cohort E and cohort B. **(A-D)** Statistical analysis assessed by Mann-Whitney rank test at each time point. Distribution of data points is determined by boxplot with median and 25^th^-75^th^ percentiles; whiskers are drawn from 1.5 times the IQR. Significant results are shown, where: *p ≤ 0.05, **p ≤ 0.01, ***p ≤ 0.001, ****p ≤ 0.0001.

S4 Fig. Influence of co-vaccination on YFV E-specific T cell responses upon vaccination in the same or different upper arms.

**(A)** Frequency of YFV E-specific CD4+ T cells compared between cohort A1 and A2. **(B)** Frequency of YFV E-specific CD4+ T cells compared between cohort B1 and B2. **(C)** Frequency of YFV E-specific CD8+ T cells compared between cohort A1 and A2. **(D)** Frequency of YFV E-specific CD8+ T cells compared between cohort B1 and B2. **(A-D)** Statistical analysis assessed by Mann-Whitney rank test at each time point. Distribution of data points is determined by boxplot with median and 25^th^-75^th^ percentiles; whiskers are drawn from 1.5 times the IQR. Significant results are shown, where: *p ≤ 0.05, **p ≤ 0.01, ***p ≤ 0.001, ****p ≤ 0.0001.

S5 Fig. Distribution of antigen-specific T cell memory phenotypes and Tfh cells.

**(A-E)** Frequencies of TEM, TCM, TEMRA, naive and Tfh of CD69+CD40L+ among CD4+ T cells for cohort C, A, D, B and E, respectively. Frequencies represents proportions of the total indicated populations. **(F-J)** Frequencies of TEM, TCM, TEMRA, and naive of CD69+41BB+ among CD8+ T cells for cohort C, A, D, B and E, respectively. Frequencies represents proportions of the total CD69+41BB+ among CD8+ T cell population.

S6 Fig. Crossreactive C-specific T cell responses in the vaccine cohorts.

**(A-B)** Frequency of TBEV, JEV, and YFV C-specific CD4+ T cells and **(C-D)** TBEV, JEV, and YFV C-specific CD8+ T cells in study participants from all cohorts expressed as fold change relative to day 0. Line plots show median and 95% confidence interval (shaded area).

S7 Fig. Crossreactive NS5-specific T cell responses in the vaccine cohorts.

**(A-B)** Frequency of TBEV, JEV, and YFV (NS5-specific CD4+ T cells and **(C-D)** TBEV, JEV, and YFV NS5-specific CD8+ T cells in study participants from all cohorts expressed as fold change relative to day 0. Line plots show median and 95% confidence interval (shaded area).

S8 Fig. Comparisons of cross-reactive E-specific CD4+ T cell responses in the vaccine cohorts.

**(A-B)** Frequency of TBEV E-specific CD4+ T cells in **(A)** cohorts C, D and E and **(B)** in cohorts A and B, expressed as fold change relative to day 0. **(A-B)** Statistical analysis assessed by Kruskal-Wallis’s and post hoc Dunn’s tests with Bonferroni correction. Distribution of data points is determined by boxplot with median and 25^th^-75^th^ percentiles; whiskers are drawn from 1.5 times the IQR. Significant results are shown, where: *p ≤ 0.05, **p ≤ 0.01, ***p ≤ 0.001, ****p ≤ 0.0001.

S9. Fig. Comparisons of cross-reactive E-specific CD8+ T cell responses in the vaccine cohorts.

**(A-B)** Frequency of TBEV E-specific CD8+ T cells in **(A)** cohorts C, D and E and **(B)** in cohorts A and B, expressed as fold change relative to day 0. **(A-B)** Statistical analysis assessed by Kruskal-Wallis’s and post hoc Dunn’s tests with Bonferroni correction. Distribution of data points is determined by boxplot with median and 25^th^-75^th^ percentiles; whiskers are drawn from 1.5 times the IQR. Significant results are shown, where: *p ≤ 0.05, **p ≤ 0.01, ***p ≤ 0.001, ****p ≤ 0.0001.

S10. Fig. Comparisons of cross-reactive C-specific CD4+ T cell responses in the vaccine cohorts.

**(A-B)** Frequency of TBEV C-specific CD4+ T cells in **(A)** cohorts C, D and E and **(B)** in cohorts A and B, expressed as fold change relative to day 0. **(A-B)** Statistical analysis assessed by Kruskal-Wallis’s and post hoc Dunn’s tests with Bonferroni correction.

Distribution of data points is determined by boxplot with median and 25^th^-75^th^ percentiles; whiskers are drawn from 1.5 times the IQR. Significant results are shown, where: *p ≤ 0.05, **p ≤ 0.01, ***p ≤ 0.001, ****p ≤ 0.0001.

S11. Fig. Comparisons of cross-reactive C-specific CD8+ T cell responses in the vaccine cohorts.

**(A-B)** Frequency of TBEV C-specific CD8+ T cells in **(A)** cohorts C, D and E and **(B)** in cohorts A and B, expressed as fold change relative to day 0. **(A-B)** Statistical analysis assessed by Kruskal-Wallis’s and post hoc Dunn’s tests with Bonferroni correction. Distribution of data points is determined by boxplot with median and 25^th^-75^th^ percentiles; whiskers are drawn from 1.5 times the IQR. Significant results are shown, where: *p ≤ 0.05, **p ≤ 0.01, ***p ≤ 0.001, ****p ≤ 0.0001.

S12. Fig. Comparisons of cross-reactive NS5-specific CD4+ T cell responses in the vaccine cohorts.

**(A-B)** Frequency of TBEV NS5-specific CD4+ T cells in **(A)** cohorts C, D and E and **(B)** in cohorts A and B, expressed as fold change relative to day 0. **(A-B)** Statistical analysis assessed by Kruskal-Wallis’s and post hoc Dunn’s tests with Bonferroni correction.

Distribution of data points is determined by boxplot with median and 25^th^-75^th^ percentiles; whiskers are drawn from 1.5 times the IQR. Significant results are shown, where: *p ≤ 0.05, **p ≤ 0.01, ***p ≤ 0.001, ****p ≤ 0.0001.

S13. Fig. Comparisons of cross-reactive NS5-specific CD8+ T cell responses in the vaccine cohorts.

**(A-B)** Frequency of TBEV NS5-specific CD8+ T cells in **(A)** cohorts C, D and E and **(B)** in cohorts A and B, expressed as fold change relative to day 0. **(A-B)** Statistical analysis assessed by Kruskal-Wallis’s and post hoc Dunn’s tests with Bonferroni correction. Distribution of data points is determined by boxplot with median and 25^th^-75^th^ percentiles; whiskers are drawn from 1.5 times the IQR. Significant results are shown, where: *p ≤ 0.05, **p ≤ 0.01, ***p ≤ 0.001, ****p ≤ 0.0001.

S14 Fig. YFV vaccine-induced T cell response shows limited cross-reactivity with ZIKV antigens.

**(A)** Sequence alignments between TBEV, JEV, YFV and ZIKV. Heatmaps are annotated according to percent of amino acid sequence similarities. **(B)** Frequency of ZIKV E, C- and NS5-specific CD4+ T cells in cohort A expressed as fold change relative to day 0. **(C)** Frequency of ZIKV E, C- and NS5-specific CD4+ T cells in cohort B expressed as fold change relative to day 0. **(D)** Frequency of ZIKV E, C- and NS5-specific CD4+ T cells in cohort E expressed as fold change relative to day 0. **(E)** Frequency of ZIKV E, C- and NS5- specific CD8+ T cells in cohort A expressed as fold change relative to day 0. **(F)** Frequency of ZIKV E, C- and NS5-specific CD8+ T cells in cohort B expressed as fold change relative to day 0. **(G)** Frequency of ZIKV E, C- and NS5-specific CD8+ T cells in cohort E expressed as fold change relative to day 0. **(B-G)** Line plots show median and 95% confidence interval (shaded area).

